# Novel manifestations of immune dysregulation and granule defects in gray platelet syndrome

**DOI:** 10.1101/2020.03.23.20041467

**Authors:** Matthew C Sims, Louisa Mayer, Janine H Collins, Tadbir K Bariana, Karyn Megy, Cecile Lavenu-Bombled, Denis Seyres, Laxmikanth Kollipara, Frances S Burden, Daniel Greene, Dave Lee, Antonio Rodriguez-Romera, Marie-Christine Alessi, William J Astle, Wadie F Bahou, Loredana Bury, Elizabeth Chalmers, Rachael Da Silva, Erica De Candia, Sri V V Deevi, Samantha Farrow, Keith Gomez, Luigi Grassi, Andreas Greinacher, Paolo Gresele, Dan Hart, Marie-Françoise Hurtaud, Anne M Kelly, Ron Kerr, Sandra Le Quellec, Thierry Leblanc, Eva B Leinøe, Rutendo Mapeta, Harriet McKin-ney, Alan D Michelson, Sara Morais, Diane Nugent, Sofia Papadia, Soo J Park, John Pasi, Gian Marco Podda, Man-Chiu Poon, Rachel Reed, Mallika Sekhar, Hanna Shalev, Suthesh Sivapalaratnam, Orna Steinberg-Shemer, Jonathan C Stephens, Robert C Tait, Ernest Turro, John K M Wu, Barbara Zieger, NIHR BioResource, Taco W Kuijpers, Anthony D Whetton, Albert Sickmann, Kathleen Freson, Kate Downes, Wendy N Erber, Mattia Frontini, Paquita Nurden, Willem H Ouwehand, Remi Favier, Jose A Guerrero

## Abstract

Gray platelet syndrome (GPS) is a rare recessive disorder caused by variants in *NBEAL2* and characterized by bleeding symptoms, the absence of platelet ɑ-granules, splenomegaly and bone marrow (BM) fibrosis. Due to its rarity, it has been difficult to fully understand the pathogenic processes that lead to these clinical sequelae. To discern the spectrum of pathological features, we performed a detailed clinical genotypic and phenotypic study of 47 GPS patients. We identified 33 new causal variants in *NBEAL2*. Our GPS patient cohort exhibited known phenotypes, including macrothrombocytopenia, BM fibrosis, megakaryocyte emperipolesis of neutrophils, splenomegaly, and elevated serum vitamin B12 levels. We also observed novel clinical phenotypes; these include reduced leukocyte counts and increased presence of autoimmune disease and positive autoantibodies. There were widespread differences in the transcriptome and proteome of GPS platelets, neutrophils, monocytes, and CD4-lymphocytes. Proteins less abundant in these cells were enriched for constituents of granules, supporting a role for Nbeal2 in the function of these organelles across a wide range of blood cells. Proteomic analysis of GPS plasma showed increased levels of proteins associated with inflammation and immune response. One quarter of plasma proteins increased in GPS are known to be synthesized outside of hematopoietic cells, predominantly in the liver. In summary, our data demonstrate that, in addition to the well-described platelet defects in GPS, there are also immune defects. The abnormal immune cells may be the drivers of systemic abnormalities, such as autoimmune disease.

## Introduction

The discovery that biallelic variants in *NBEAL2* cause gray platelet syndrome (GPS) has been a key advance in hematology^1-3^ and has improved our understanding of platelet and granule biology^4,5^. The *Nbeal2*^*-/-*^ mouse phenocopies the features of GPS, including bleeding, thrombocytopenia, absence of platelet alpha (ɑ)-granules, splenomegaly, and bone marrow (BM) fibrosis^6-8^. In addition to the known defects of the megakaryocytic-platelet axis, we and others have reported the susceptibility of *Nbeal2*^*-/-*^ mice to bacterial and viral infection, which is implicated in increased organ damage, lower survival rates, and longer time to recovery upon infection^9,10^. These observations have been attributed to defects of secretory granules in natural killer cells and neutrophils. Additionally, it has been shown that mast cells of *Nbeal2*^*-/-*^ mice are deficient in storage vesicles^11^ and that monocytes have reduced granularity^9^. Together with our observation that *Nbeal2*^*-/*-^ megakaryocytes (MKs) have a pro-inflammatory profile^7^, these findings highlight that Nbeal2 is important for normal granule function in MKs, platelets, and a variety of myeloid and lymphoid cells in mice. However, it has been unclear whether the immune-cell defects seen in the murine model are relevant to the pathophysiology of GPS. Small series of GPS patients have reported recurrent infection^12-14^ and atypical presentations of autoimmune lymphoproliferative syndrome (ALPS)^15,16^. Neutrophil ultrastructure has been evaluated in several GPS patients with conflicting results: some reports have shown that neutrophil granules are preserved^17,18^, whilst others have noted their absence^12,13^. Given the paucity of published studies on GPS patients with confirmed causal variants in *NBEAL2*, we established an international collaboration to systematically evaluate clinical and laboratory phenotypes in a large collection of patients. We focused on features related to immunity and followed this up with a detailed molecular characterization of plasma, platelets, and three different leukocyte populations in a subset of the patients using both protein mass spectrometry (MS) and RNA sequencing (RNA-seq).

## Methods

This section contains a short description of the methods we have used. Please refer to the Supplemental (Supp.) Data for further details.

### Enrollment

All study participants provided written informed consent. The majority of participants enrolled into UK or French studies, all of which were approved by research ethics committees (UK: REC 13/EE/0325, REC 10/H0304/65, REC 10/H0304/66; France: INSERM RBM-014). The remaining participants consented using ethics procedures approved at the recruiting center.

### Sequencing and variant interpretation

DNA was isolated and sequenced using the ThromboGenomics next generation sequencing platform, long range PCR, or whole genome sequencing^1,19-21^. Variant selection was based on the assessment of the center of enrollment or the criteria used by the Thrombogenomics consortium^19^. Pathogenicity classification of variants was carried out in our established multi-disciplinary meetings and only pathogenic variants (PV), likely pathogenic variants (LPV), and variants of unknown significance (VUS) in *NBEAL2* were selected for further analysis^19,22^. We only included patients with causal variants in both alleles of *NBEAL2*.

### Phenotyping of patients

Clinical information and the results of laboratory investigations of the patients were obtained by enrolling physicians. This information was used to record human phenotype ontology (HPO) terms for each participant^23^.

### BM biopsies

A BM biopsy was performed on 23 GPS patients. BM fibrosis was defined by a reticulin grade ≥ 2, diagnostic by WHO criteria^24^, as determined by the host institution. Five BMs were also reviewed centrally. Quantification of emperipolesis, the presence of intact neutrophils in the cytoplasm of MKs, was performed by manual counting on three samples.

### Enhanced cellular phenotyping

Five of the GPS patients and five age-matched controls attended the NHS Blood and Transplant Centre in Cambridge (UK) for a more complete evaluation under standardized conditions. Following fasting, a complete blood count (CBC) was performed on an EDTA anticoagulated blood sample using a Sysmex XN-1000 analyzer. Additional measured blood cell parameters, not normally included in CBC reports including forward scatter (FSC) and side scatter (SSC), were analyzed; FSC indicates cell volume and SSC reflects the complexity of cellular contents, including granularity. In addition, platelets, neutrophils, monocytes, and CD4-lymphocytes were isolated from citrate anticoagulated blood using established protocols^25^. For each cell type, RNA was extracted and sequenced, and the lysate was analyzed by protein MS.

### Plasma proteomics

Plasma was obtained from EDTA anticoagulated blood of 11 GPS patients and 13 age-matched controls then analyzed using protein MS.

### Statistics

The Fisher’s exact test was used for the majority of statistical analyses, unless stated otherwise.

### Data sharing statements

RNA-seq data has been deposited in the European Genome-phenome Archive under study accession number ID EGAD00001005950. All newly identified *NBEAL2* variants have been deposited in ClinVar. Cell-type specific and plasma proteomics results have been deposited to the ProteomeXchange Consortium via the PRIDE repository^26^ at EMBL-EBI with the dataset identifiers PXD016366 and PXD017227, respectively.

## Results

### A wide spectrum of *NBEAL2* variants cause GPS

Forty-seven patients from 38 genetically independent pedigrees were enrolled at 21 hospitals in 11 countries. We identified a total of 72 causal variants in *NBEAL2*, 33 of which are reported for the first time in this study (Fig. 1A; Supp. Table 1). All variants were rare with a minor allele frequency in the Genome Aggregation Database of 0 (i.e. absent) to 0.000029 (1 in 34,500)^27^. Of the total variants identified, 43% were inherited in homozygosity and 57% in compound heterozygosity. Fifty-eight variants were unique, 86% of which were classified as PV or LPV (Fig. 1B). Missense variants accounted for just under half of all causal variants identified, with the remaining ones being categorized (in descending order of frequency) as frameshift, stop gained, splice, and short insertion/deletion (indel) variants. We mapped the position of the missense variants onto the five known domains of Nbeal2 and determined the evolutionary conservation score of the affected residue using ConSurf ^28^ (Fig. 1C). Missense variants were enriched (*P =* 1.85 × 10^−5^) in the highly conserved BEACH do-main, consistent with a suggested key role of this domain in the function of Nbeal2^5,29^.

**Figure 1.**
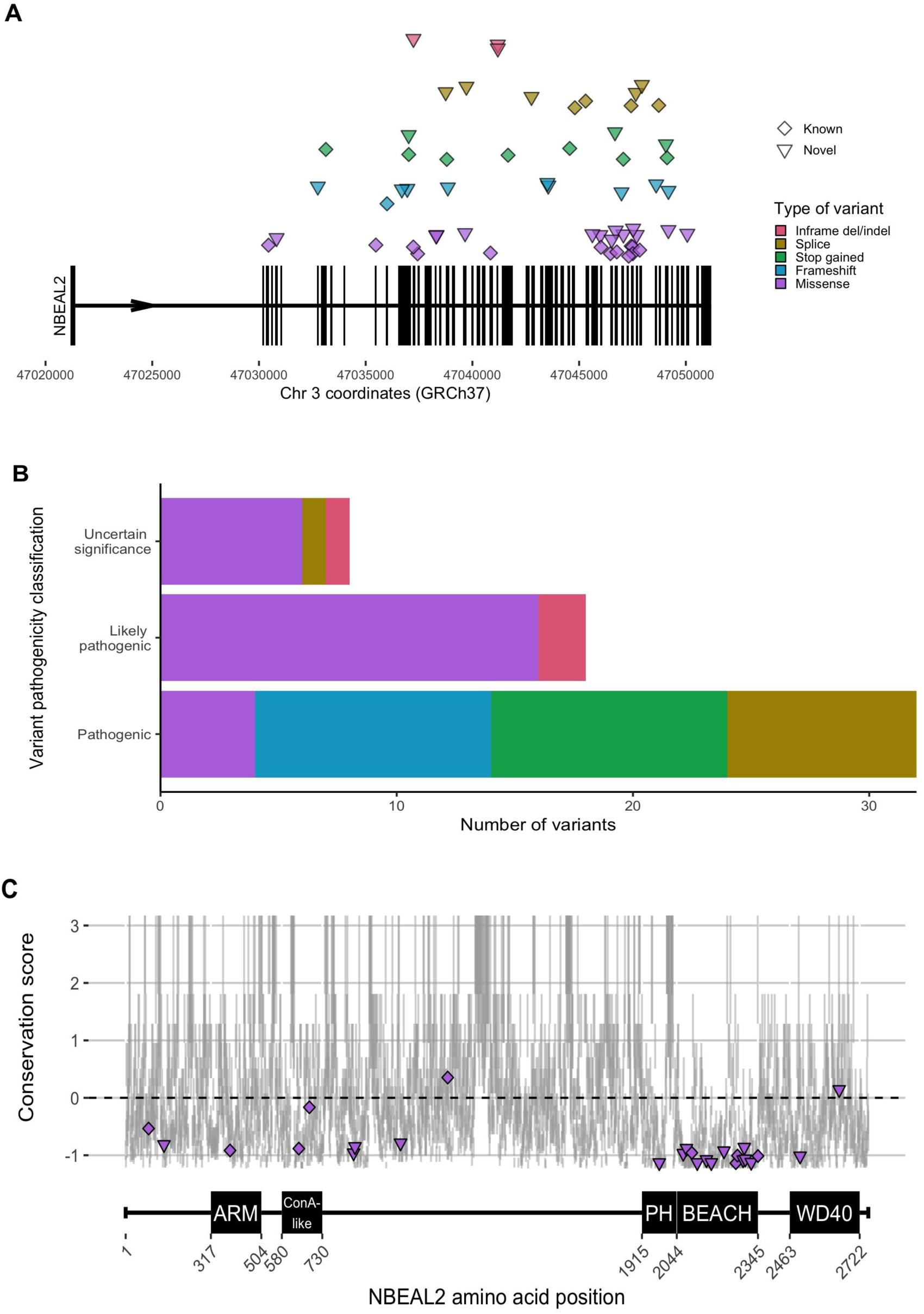
*NBEAL2* variants identified in GPS patients participating in this study. **A)** Position of the 58 unique variants relative to *NBEAL2* on chromosome 3 (Chr 3) using Genome Reference Consortium human genome build 37 (GRCh37). Vertical black bars represent exons. The color and shape used to represent variants in the legend applies throughout the figure. **B)** The frequency of unique variants classified by pathogenicity, according to the American College of Medical Genetics and Genomics guidelines^22^. **C)** The location of missense variants compared to the amino acid sequence of Nbeal2 and known functional domains (on the x-axis). The y-axis position of the variant is its estimated evolutionary conservation score. A positive score corresponds to lower conservation and a negative score represents higher conservation. Vertical gray lines represent the range of conservation scores of the 50% confidence interval: the bottom and top of the bar are the 25th and 75th percentile, respectively, of the inferred evolutionary rate distribution.

We then mapped the 14 BEACH domain missense variants to the crystal structure of another BEACH domain containing protein, Nbea^30^. Five variants occurred in the hydrophobic core of the domain, a portion of which interacts with the neighboring PH domain, a 129-residue domain with a neutral conservation score (0.002)^28^. However, we identified a novel missense variant, R1979W, in a portion of the PH domain which is highly conserved; the position of this variant corresponds to Nbea R2208, which has been shown to be functionally relevant for the PH-BEACH interface^30^. To our knowledge, this is the first GPS-causing variant to be described in the PH domain.

### Bleeding and platelet phenotypes in GPS

Patients recruited into the study were between the ages of 6 and 70 years with a median of 35. The median age at presentation was 11.5 years, but this varied between 2 months and 67 years. All patients were thrombocytopenic (Fig. 2A; Supp. Fig. 2.1; Supp. Table 2). Consistent with previous reports^17,31,32^, a wide spectrum of bleeding symptoms were reported (Supp. Fig. 2.1), ranging from subcutaneous to intracranial hemorrhage, which in one case was fatal. Five patients were notable for their lack of a bleeding diathesis. Alpha-granule deficiency was noted in the platelets of all cases assessed by electron or light microscopy (Fig. 2A).

**Figure 2.**
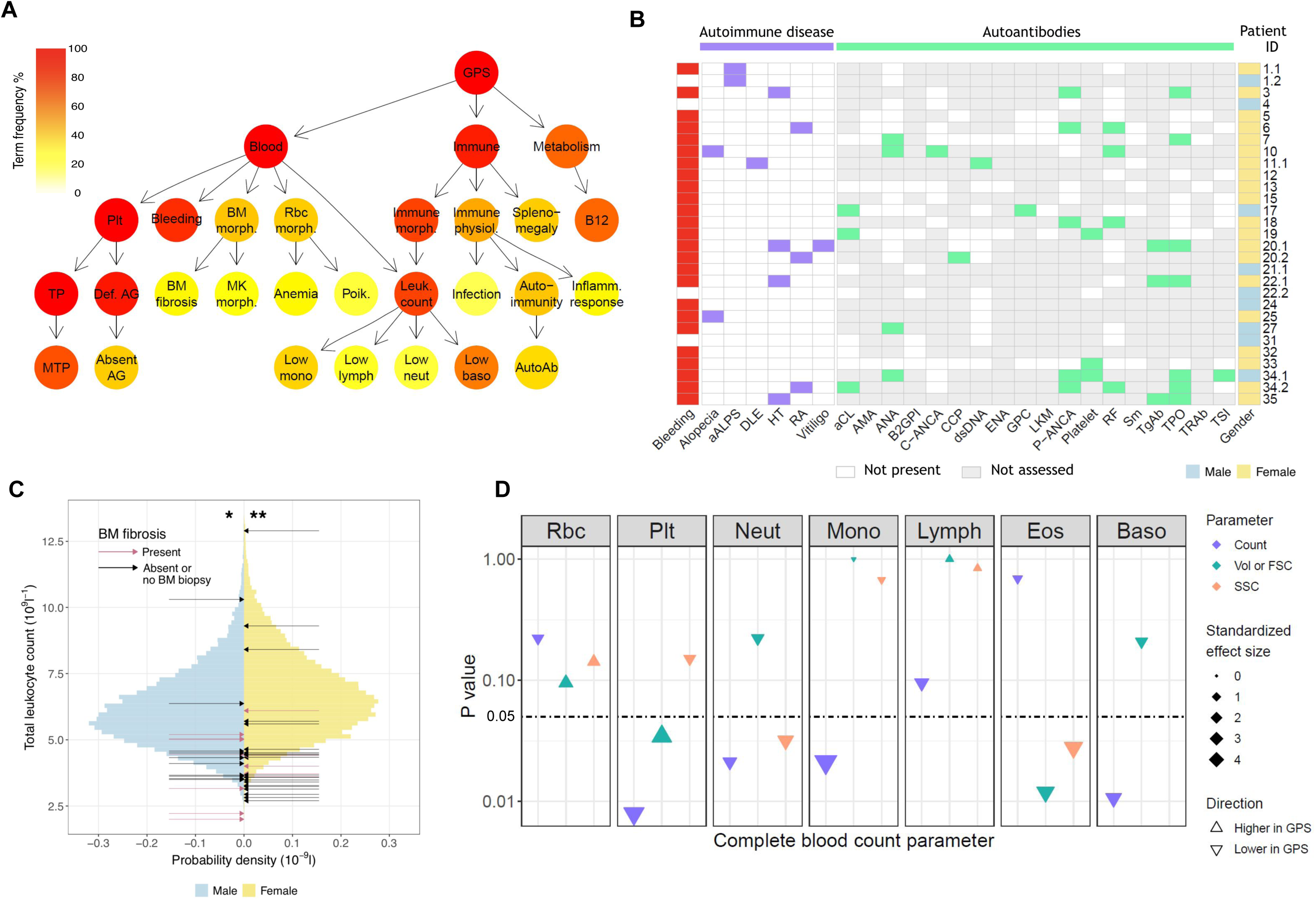
Novel clinical phenotypes. **A)** Summary Human Phenotype Ontology (HPO) tree, showing the three most frequent HPO organ systems represented in the 47 GPS patients: Blood (“Abnormality of blood and blood-forming tissues”), Immune (“Abnormality of the immune system”), and Metabolism (“Abnormality of metabolism/homeostasis”). HPO terms affecting eight or more patients are shown except terms associated with “Bleeding”, which are displayed in Supp. Fig. 2.1. HPO term labels: Plt, “Abnormal thrombocyte morphology”; Bleeding, “Abnormal bleeding”; BM morph, “Abnormality of bone marrow cell morphology”; Rbc morph., “Abnormal erythrocyte morphology”; Immune morph., “Abnormal immune system morphology”; Immune physiol., “Abnormality of immune system physiology”; Splenomegaly, “Splenomegaly”; B12, “Abnormal vitamin B12 level”; TP, “Thrombocytopenia”; Def AG, “Abnormal number of alpha granules”; BM fibrosis, “Myelofibrosis”; MK morph., “Abnormal megakaryocyte morphology”; Anemia, “Anemia”; Poik., “Poikilocytosis”; Leuk. count, “Abnormal leukocyte count”; Infection, “Recurrent infections”; Autoimmunity, “Autoimmunity”; Inflamm. response, “Increased inflammatory response”; MTP, “Macrothrombocytopenia”; Absent AG, “Absence of alpha granules”; Low mono, “Monocytopenia”; Low lymph, “Lymphopenia”; Low neut, “Neutropenia”; Low baso, “Decreased basophil count”; AutoAb, “Autoimmune antibody positivity”. **B)** Representation of autoimmune disease, results of autoantibody tests and presence of bleeding symptoms for 29 GPS patients (labelled with Patient ID). Autoantibodies tested in at least three patients are included. Abbreviations: aALPS, atypical autoimmune lymphoproliferative syndrome; DLE, discoid lupus erythematosus; HT, hashimoto’s thyroiditis; RA, rheumatoid arthritis; and autoantibodies against: aCL, cardiolipin; AMA, mitochondria; ANA, nuclear antigen; B2GPI, beta-2 glycoprotein I; C-ANCA, neutrophil cytoplasmic antigen; CCP, cyclic citrullinated peptide; dsDNA, double stranded DNA; ENA, extractable nuclear antigen; GPC, gastric parietal cell; LKM, liver-kidney microsome; P-ANCA, neutrophil perinuclear antigen; RF, rheumatoid factor; Sm, small nuclear ribonucleoprotein; TgAb, thyroglobulin; TPO, thyroperoxidase; TRAb, thyroid stimulating hormone receptor; TSI, thyroid stimulating immunoglobulin. **C)** Histogram showing the total leukocyte count of 45,032 blood donors in the INTERVAL study^34^ stratified by sex upon which the results of the GPS patients are represented by arrows. The median total leukocyte count of both male and female GPS patients was significantly (*P* = 3×10^−3^ and 4×10^−4^, respectively) lower than INTERVAL participants using a one-sample Wilcoxon signed-rank test. **D)** Complete blood count (CBC) results for five GPS patients versus five controls. The data point for each cell type and CBC parameter shows the absolute standardized effect size and directionality. On the y-axis (log10 scale), the *P* value (calculated using the Mann-Whitney U test) is shown; the horizontal dotted line represents *P* < 0.05. Volume (Vol) or forward scatter (FSC) refers to the following measurements: mean cell volume (MCV) for red blood cells (Rbc), platelet mean frequent volume (P-MFV) for platelets (Plt); forward scatter (FSC) for neutrophils (Neut), monocytes (Mono), lymphocytes (Lymph), eosinophils (Eos), and basophils (Baso). Side scatter (SSC) was available for all parameters except basophils.

### Raised B12 levels and stable BM fibrosis

Raised serum vitamin B12 levels (B12) were recorded for 31 out of 34 patients, expanding the previous observations made in a smaller GPS collection^17^. B12 was increased by at least 1.5 times the upper limit of the local reference range in two thirds of patients (Supp. Fig. 2.1; Supp. Table 2).

Of the BM biopsies performed in 23 patients, 13 (57%) were diagnosed with BM fibrosis. Morphological examination of the five centrally reviewed trephine samples showed preservation of trilineage hematopoiesis (Supp. Fig. 2.2). BM fibrosis was diagnosed at a median age of 28.5 years (range 10 - 52), with a median of 16 years since this diagnosis to the present time. Patient 20.3 received an allogeneic stem cell transplant at age 32 years^33^, but the rest of the patients did not require treatment for BM fibrosis during this time. Neutrophil emperipolesis by MKs was observed in a mean of 58% MKs in three GPS BM trephines compared to only 1% in three trephines from controls (Supp. Fig. 2.2).

### GPS patients have abnormalities of the immune system, including autoimmune disease affecting multiple organ systems

Coding of clinical and laboratory information by HPO terms identified phenotypes associated with ‘Abnormality of the immune system’ in 91% of GPS patients (Fig. 2A; Supp. Table 2). ‘Splenomegaly’ (determined clinically or by ultrasound) was observed in 40% of patients and did not significantly overlap with BM fibrosis (ChiSquared test, *P* = 0.26; Supp. Fig. 2.2). ‘Abnormal immune system morphology’ (81%) was driven predominantly by cytopenia of at least one leukocyte type (77%). ‘Abnormal immune system physiology’ (51%) included ‘Recurrent infec-tions’ (17%), most commonly respiratory tract infections and otitis media, and ‘Autoimmunity’ (43%), which comprised of autoimmune disease and positive serology. Twelve patients (26%) were diagnosed with an autoimmune disease with a wide spectrum of organ systems affected. These include endocrine (Hashimoto’s thyroiditis), skeletal (rheumatoid arthritis), integumentary (alopecia, discoid lupus erythematosus, and vitiligo), and the immune system (atypical ALPS) (Fig. 2B). Twenty-nine patients had autoantibody tests performed and, of these, 17 (59%) had at least one positive test with a female:male ratio of 5:1. The four most frequent positive tests (in descending order of frequency) were anti-thyroperoxidase (TPO), perinuclear antineutrophil cytoplasmic antibodies (P-ANCA), rheumatoid factor (RF), and anti-nuclear antibodies (ANA).

### Altered blood cell parameters of multiple cell lineages in GPS patients

We compared the complete and differential leukocyte counts of all 47 GPS patients against those of 45,000 healthy blood donors from the INTERVAL study^34^. The median total leukocyte count of both male and female GPS patients was significantly lower than for the INTERVAL participants (one-sample Wilcoxon signed-rank test, *P* = 3×10^−3^ and 4×10^−4^, respectively) (Fig. 2C). All median differential leukocyte counts (neutrophil, monocyte, lymphocyte, eosinophil, and basophil counts) as well as platelet counts and hemoglobin concentrations were significantly lower than the corresponding INTERVAL values (Supp. Fig. 2.1 and 2.3; Supp. Table 2). Although we found no association between total leukocyte, granulocyte, or monocyte counts with BM fibrosis (logistic regression, *P* > 0.05), platelet and lymphocyte counts were inversely associated with the latter (logistic regression, *P* < 0.05). Furthermore, there was no association between total or any of the differential leukocyte counts and splenomegaly (logistic regression, *P* > 0.05).

**Figure 3.**
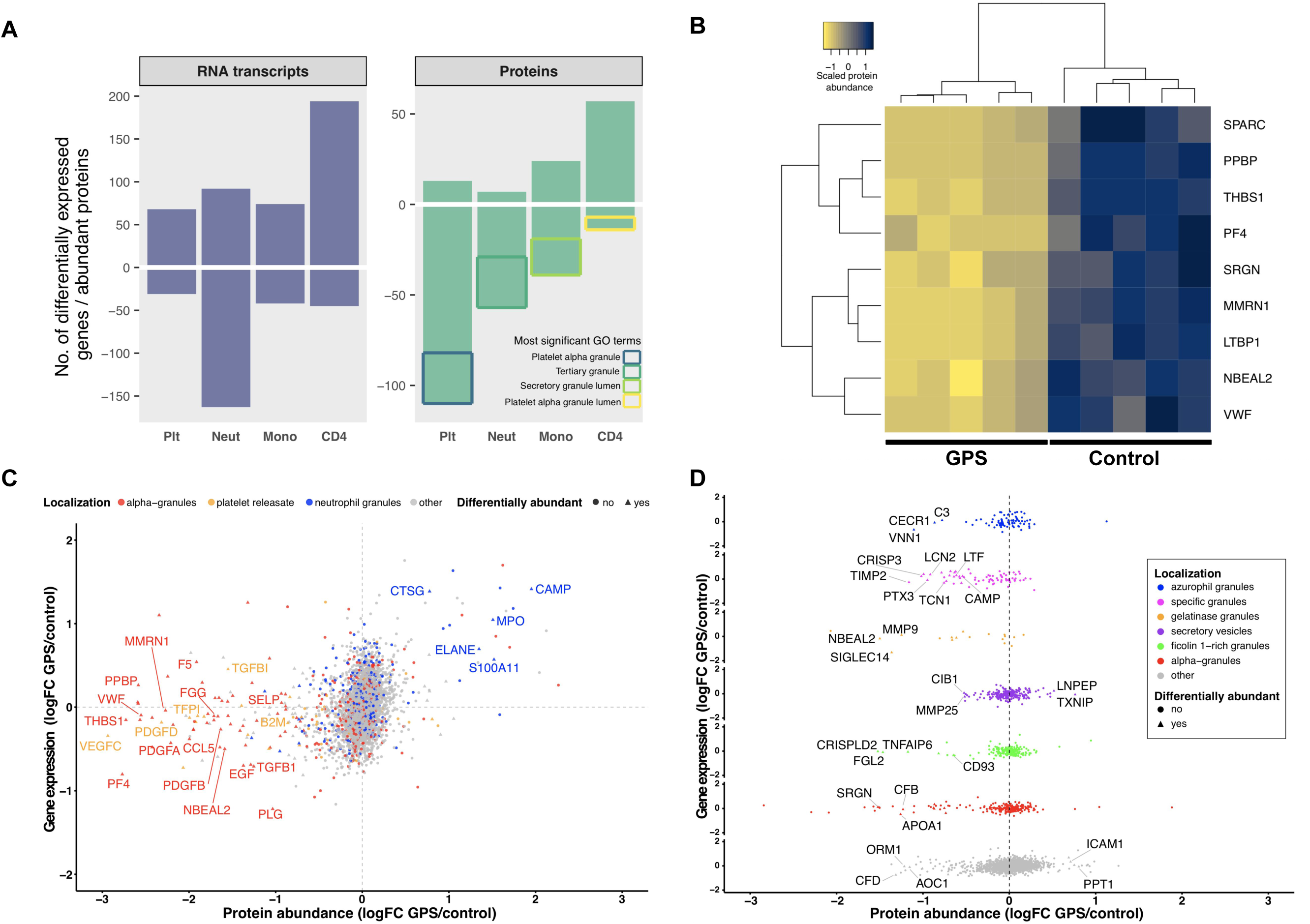
The transcriptome and proteome of platelets, neutrophils, monocytes and CD4-lymphocytes. For all panels, log2 fold change (logFC) for gene expression (by RNA-seq) and protein abundance (by MS) was determined by log2-transforming the ratio of the means of individual normalized values of the five GPS patients and five controls. Negative values represent genes/proteins that are reduced in GPS and vice-versa. The threshold for differential abundance/expression was determined by each experimental method and the corresponding analysis used (Supp. Data). **A)** Bar chart showing differentially expressed genes by RNA-seq (left) and differentially abundant proteins by MS (right). The y-axis represents the absolute number of genes or proteins determined by each method and negative numbers represent significant reduction in GPS cells and vice versa. The most significant gene ontology (GO) cellular component terms for the differentially less abundant proteins of each of the cell types is illustrated in the “Proteins” panel. **B)** For each individual, the protein abundance was calculated for each of the nine proteins that are differentially less abundant in at least three types of cells and averaged across all of the cells. The results were scaled from −1 to 1 and plotted as a dendrogram with heatmap, such that positive (dark blue) and negative (yellow) values represent higher and lower protein abundance, respectively. Hierarchical clustering was applied using the complete linkage method and dissimilarity between rows and columns was based on the Euclidean distance. Each column represents an individual GPS patient or control. **C)** The comparison of gene expression and protein abundance of platelets (GPS vs control) highlights the high proportion of “ɑ-granules” and “releasate” proteins (in red and orange, respectively), which are significantly diminished in GPS platelets at the protein level (*P* < 2.2×10-16), and also shows that proteins normally resident in neutrophil granules (blue dots) are increased in GPS platelets. **D)** The comparison of gene expression and protein abundance of neutrophils (GPS vs control) separated vertically by subcellular localization shows a significantly lower abundance of specific and gelatinase granule proteins in GPS neutrophils (*P* < 1.3×10^−8^ and < 2.2×10^−16^, respectively).

In addition, five GPS patients and five controls participated in a single center followup study where CBC measurements were carried out that included additional parameters (FSC and SSC) not available from clinical records. This analysis confirmed that the neutrophil, monocyte, and basophil counts were significantly reduced in GPS patients (Mann–Whitney U test, *P* < 0.05). Moreover, the FSC and SSC measurements showed a significant reduction in GPS eosinophil size and granularity, the latter also holding true for neutrophils (Mann–Whitney U test, *P* < 0.05; Fig. 2D).

### Genotype-phenotype analysis

To determine whether the type of causal variant influenced the severity of clinical features, we grouped patients into those with biallelic loss of function (LoF) variants (frameshift, stop gained, splice, and indel) and those with biallelic missense variants, excluding patients with compound heterozygosity for one LoF and one missense variant. There were 24 patients in the LoF group and 17 patients in the missense group. There were no significant differences in age (current and at presentation), spleen size, and CBC parameters between the groups (Welch two sample t-test and Wilcoxon rank sum test, *P* > 0.05; Supp. Table 2). There was no significant over-representation of BM fibrosis, susceptibility to infection, or autoimmune diseases in one group compared to the other (*P* > 0.05). We then grouped patients with missense variants into those with biallelic BEACH-domain variants (n = 7) and those with biallelic variants outside of the BEACH domain (n = 5), excluding compound heterozygotes carrying one BEACH and one non-BEACH variant, and repeated the above analysis using the same variables and statistical tests. Again, no significant differences could be detected between the two groups.

### Platelets and leukocytes have altered transcriptome and proteome profiles in GPS

Prompted by the propensity to autoimmune diseases and the altered CBC parameters, we set out to obtain a more comprehensive understanding of the differences in cellular phenotypes of the same five GPS patients who had additional CBC parameters measured. Platelets, neutrophils, monocytes, and CD4-lymphocytes were analyzed by RNA-seq and protein MS and results compared to those obtained for the five controls. Principal component analysis of these data clearly delineated GPS and control samples in all experiments, except for platelet RNA-seq (Supp. Fig. 3.1). Across each of the four blood cell types, the number of differentially expressed genes ranged from 95 to 255 and differentially abundant proteins from 63 to 123 (Fig. 3A; Supp. Table 3). Of the 123, 65, and 63 differentially abundant proteins in platelets, neutrophils, and monocytes, 89%, 86%, and 62% were reduced in GPS patients, respectively. These reduced proteins were enriched in gene ontology (GO) terms pertaining to cell granules and their lumens. Nine proteins, including Nbeal2, were significantly reduced in GPS patients in at least three of the blood cell types, and all are known to localize to blood cell granules (Fig. 3B).

### GPS platelets are diminished in ɑ-granule cargo but contain neutrophil granule proteins

Of the proteins differentially less abundant in GPS platelets, there was a significant over-representation of proteins known to be present in ɑ-granules and/or the platelet releasate (73 out of 110, *P* < 2.2×10^−16^; Fig. 3C; see Supp. Data for classification of protein localization). Interestingly, none of these proteins were differentially expressed in the RNA-seq analysis, suggesting that a loss of function of Nbeal2 does not affect the transcriptional output of ɑ-granule genes in megakaryocytes. We then inspected the 13 proteins differentially more abundant in GPS platelets. Five of these are archetypal neutrophil granule constituents, such as Elastase (Elane) and Myeloperoxidase (Mpo) (Fig. 3C; Supp. Fig. 3.2). Indeed, when analyzing the platelet proteomics data using a less stringent cut-off (logFC> 0.5) than that used to determine differential protein abundance (Supp. Data), we identified 54 proteins more abundant in GPS platelets with a significant over-representation of neutrophil granule constituents (n = 13, *P* = 0.02).

### GPS neutrophils and monocytes are deficient in granule proteins

To initially explore the GPS neutrophil proteome, we annotated the differentially abundant proteins by their presence or absence in granules. For known granule proteins these were further annotated by granule subtype (Supp. Data). Of the 56 differentially abundant proteins that were reduced in GPS neutrophils (Fig. 3A), 36 (65%) could be assigned to their different granule subtypes (Fig. 3D; Supp. Fig. 3.3; Supp. Table 3). Notably, only the gelatinase and specific granules were significantly over-represented (*P* < 1.3×10^−8^ and < 2.2×10^−16^, respectively). Given the reduced counts of monocytes in our GPS patients, we hypothesized that a lack of functional Nbeal2 could also influence the granularity of these cells. Although there is no authoritative publication on the monocyte granule proteome, we annotated the differentially abundant proteins as granule proteins if they overlapped with proteins detected in platelet releasate, ɑ-granules, or neutrophil granules. Of the 39 proteins reduced in GPS monocytes, 29 had a granular localization (Supp. Fig. 3.4). Similar to GPS platelets and neutrophils, the genes encoding these 29 proteins were not differentially expressed.

**Figure 4.**
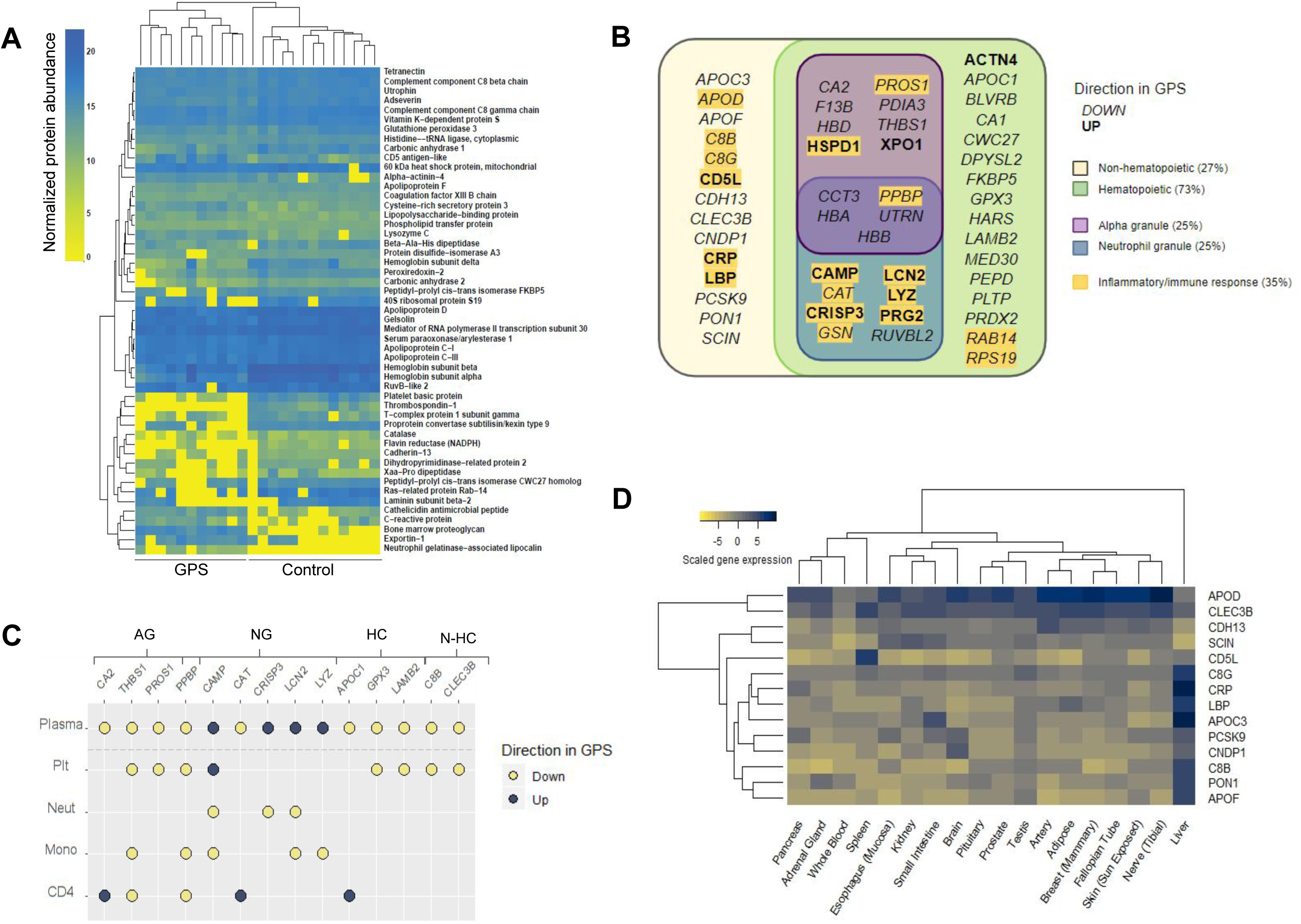
The GPS plasma proteome. **A)** A heatmap of normalized protein abundance of the 51 discriminatory proteins detected in plasma from 11 GPS patients and 13 controls which showed clear separation by unsupervised clustering. Each column represents an individual GPS patient or control. **B)** Classification of the 51 discriminatory proteins of the plasma proteome by directionality in GPS plasma, cellular origin (hematopoietic or non-hematopoietic) and subcellular localization (platelet alpha or neutrophil granules). Proteins involved in inflammatory or immune responses are highlighted. **C)** A dot plot representing the 14 discriminatory plasma proteins that are also differentially abundant proteins in at least one of the cell types evaluated by mass spectrometry (platelets [Plt], neutrophils [Neut], monocytes [Mono], and CD4-lymphocytes [CD4]). Each data point represents a protein and is color-coded by its directionality in the respective proteome. Absence of a protein in an explicit cell-specific proteome denotes a protein that was determined not to be differentially abundant. Proteins are annotated by granule localization (AG = platelet alpha granule, NG = neutrophil granule) or, for proteins not localized in granules, gene expression in hematopoietic cells (HC) or non-hematopoietic cells (N-HC) in the BLUEPRINT consortium atlas^37^. **D)** A heatmap showing the gene ex-pression of the 14 discriminatory plasma proteins of non-hematopoietic origin in the Genotype-Tissue Expression Program^38,39^. Scaled gene expression represents log2 transformation of transcripts per million base pairs mapped.

### GPS CD4-lymphocytes up-regulate markers of immune responses

Given the associations between CD4-lymphocytes and autoimmune disorders^35^, we also included these cells for evaluation by RNA-seq and protein MS. In contrast to the other three cell types analyzed, the majority of differentially abundant proteins in GPS CD4-lymphocytes, 57 out of 71, were increased and there was a correlation with gene expression (Supp. Fig. 3.4; Supp. Table 3). Among these proteins/genes differentially more abundant and more highly expressed, there was an over-representation of proteins involved in immunomodulatory functions, e.g. Bruton Tyrosine kinase (Btk), the α-chain of the receptor for the Fc domain of IgE (Fcer1a), and Bone marrow stromal cell antigen-1 (Bst1).

### The GPS plasma proteome has a pro-inflammatory and hepatic signature

We postulated that the broad granular abnormalities in the blood cells evaluated might lead to systemic changes in the blood circulation. To this end, we analyzed the plasma of 11 patients and 13 controls using protein MS. First, an unsupervised random forest analysis of the normalized concentration of plasma proteins segregated patient and control samples and demonstrated that there were 51 discriminatory proteins (Fig. 4A; Supp. Table 3). The analysis showed that 11 and 40 discriminatory proteins had higher and lower concentrations, respectively, in patients. GO enrichment analysis highlighted the presence of terms associated with inflammation and immune response, particularly in the top-ranked terms for the 11 proteins more abundant in GPS plasma (Fig. 4B). We then overlapped the discriminatory plasma proteins with the proteome of the four cell types analyzed. Fourteen of the discriminatory plasma proteins were differentially abundant in the proteome of at least one cell type, but the pattern of directionality differed by protein (Fig. 4C). Interestingly, proteins that were less abundant in the GPS platelet proteome were also less abundant in GPS plasma, including proteins localized to ɑ-granules. In contrast, all of the four overlapping proteins with higher levels in plasma are present in neutrophil granules. Of these, Cathelicidin antimicrobial peptide (Camp), Cysteine-rich secretory protein 3 (Crisp3), and Neutrophil gelatinase-associated lipocalin (Lcn2) were all differentially less abundant in GPS neutrophils, and all localize to the neutrophil specific granules^36^.

Cross-referencing the discriminatory plasma proteins with the BLUEPRINT consortium gene expression data^37^ revealed that 14 of the discriminatory plasma proteins are not known to be expressed in hematopoietic cells (Fig. 4B; Supp. Fig. 4.1). We then analyzed the gene expression of these 14 proteins in the Genotype-Tissue Expression (GTEx) database^38,39^, which demonstrated that nine of these non-hematopoietic proteins are predominantly synthesized by liver-residing cells. These include known acute phase reactants such as C-reactive protein (Crp) and Lipopolysaccharide-binding protein (Lbp) (Fig. 4D), both of which had increased levels in the plasma of GPS patients.

## Discussion

Through our international collaboration, we have brought together the largest collection of GPS patients to date. We report on 58 unique causal *NBEAL2* variants, which are significantly enriched within the BEACH domain. Thirty-three variants were newly identified, including one in the PH domain, which we predict is located at the PH-BEACH interface. By comprehensively re-assessing established phenotypes of GPS we can confirm that there is marked heterogeneity of bleeding symptoms, a near universal increase in B12, and BM neutrophil emperipolesis^17,31,40^. Furthermore, our data suggest that splenomegaly and BM fibrosis each occur in around half of patients, but without significant overlap. Importantly, in the majority of patients, BM fibrosis appears to be non-progressive and is likely to be a reactive and secondary phenomenon that does not generally require intervention. We demonstrate that the median count for all leukocyte types is reduced in GPS and show that neutrophil granularity, as assessed by SSC, is reduced in GPS. We also show, for the first time, that eosinophil SSC is diminished, suggesting a potential role for Nbeal2 in the granule biology of these cells.

Immune dysregulation in patients was common with over half having detectable autoantibodies and one quarter clinically manifesting autoimmune disease. Comparison with HPO terms in a control collection of 428 patients with suspected inherited thrombocytopenia^21^, revealed autoimmunity in 3% of the patients, suggesting an over-representation of this phenotype in GPS. We observed a wide spectrum of immune pathologies including newly-described associations with rheumatoid arthritis, alope-cia, and skin-related autoimmune disorders. In addition, we extend a previous single case report of Hashimoto’s thyroiditis^12^ to four GPS patients.

Almost 20% of the patients had recurrent infections corroborating our observations in *Nbeal2*^-/-^ mice^10^. This wide spectrum of autoimmune diseases and immunodeficiency in the context of granule defects are also the consequence of Mendelian disorders of other BEACH-domain containing genes, such as *LYST, LRBA*, and *WDFY4*^29^, and highlights the disruptive effect of etiological variants in *NBEAL2* on a variety of immune cells. This was demonstrated in the RNA-seq and protein MS analysis of platelets, neutrophils, monocytes, and CD4-lymphocytes, where there were widespread differences in the transcriptome and proteome of all cells evaluated; this is in stark contrast to the isolated platelet defect seen in our recent description of *IKZF5*-related thrombocytopenia, a disorder also accompanied by reduced α-granularity^25^.

As expected, GPS platelets are markedly diminished in proteins known to localize to ɑ-granules. Unexpectedly, we observed the ectopic presence of proteins normally resident in neutrophil granules in GPS platelets. This suggests that the consequence of neutrophil emperipolesis by MKs is not only a BM phenomenon, but has ramifications for circulating platelets. Indeed, the transfer of membrane between MKs and neutrophils during emperipolesis has been recently reported^41^ and a recent functional genomic study has suggested the association of Mpo, known to localize to neutrophil specific granules, with mean platelet volume highlighting the cross-talk between neutrophils and platelets^42^. Our proteomic dataset strongly supports the reduction of neutrophil specific granules in GPS, an observation that has previously been con-tested^12,13,18^, and shows for the first time that gelatinase granule constituents are also depleted from GPS neutrophils. Our results also show a significant depletion of granule proteins in the monocytes of GPS patients, which aligns with the reduced granularity of monocytes reported in *Nbeal2*^*-/-*^ mice^9^ and provides evidence for the function of Nbeal2 in monocyte granules.

The cytoplasm of CD4-lymphocytes is predominantly occupied by their nucleus and the majority of these cells do not contain secretory granules, which may provide an explanation for the underrepresentation of granule proteins in the differential transcriptome and proteome of GPS CD4-lymphocytes. However, GPS CD4-lymphocytes showed the strongest correlation between the components of the transcriptome and the proteome with a significant up-regulation of multiple transcripts and their corresponding proteins. This up-regulation is enriched in GO terms related to immune responses, e.g. *FCER1A* and its encoded high affinity receptor for IgE (FceR1a), which had a high logFC in both the GPS transcriptome and proteome. Likewise, *BTK*, which encodes Btk and is critical for B-cell development, was both more highly-expressed and its protein differentially more abundant in GPS CD4-lymphocytes compared to controls. Interestingly, recent studies suggest that Btk also plays an important role in the regulation of T cells and its increased levels could contribute to the widespread autoimmunity observed in our GPS patients^43,44^.

Although we observed a depletion of granule proteins in GPS neutrophils and monocytes, some granule proteins, e.g. Camp, which has a function in innate immunity, as well as other proteins related to pathogen defense and acute phase response, were in-creased in the patient plasma. Furthermore, over a quarter of the discriminatory plasma proteins are not known to be synthesized by blood cells, the majority of which are produced in liver-residing cells. Possible mechanisms to explain this observation in GPS patients include the systemic activation of the liver by pro-inflammatory myeloid granule proteins and/or the abnormal infiltration of atypical blood cells into the liver.

In summary, our study identifies novel features of GPS including reduced leukocyte counts, autoimmune diatheses, hypogranularity of myeloid cells, and a pro-inflammatory plasma proteome. In a recent genome-wide association study (GWAS) for CBC parameters in ∼500,000 individuals in the UK Biobank, a variant associated with platelet distribution width was identified in close proximity to the transcription start site of *NBEAL2* (Vuckovic et al., manuscript submitted January 2020). This observation supports the notion that *NBEAL2* variants present on one allele have biological consequences for platelet morphology and is supported by the findings that GPS family members carrying one GPS-causing variant exhibit partial ɑ-granule deficiency and platelet macrocytosis^45^. Therefore, carriers of pathogenic *NBEAL2* variants may present with late onset pathologies reminiscent of GPS, an assumption which can be explored in the future with the ever-increasing number of individuals participating in population GWAS.

## Data Availability

RNA-seq data has been deposited in the European Genome-phenome Archive under study accession number ID EGAD00001005950. All newly identified NBEAL2 variants have been deposited in ClinVar. Cell-type specific and plasma proteomics results have been deposited to the ProteomeXchange Consortium via the PRIDE repository at EMBL-EBI with the dataset identifiers PXD016366 and PXD017227, respectively.

https://www.ebi.ac.uk/ega/datasets/EGAD00000000021

https://www.ebi.ac.uk/pride/

## Acknowledgements

We would like to thank Rachel Linger (NIHR BioResource) for assistance with recruitment of patients and healthy donors, Amy Frary and Carly Kempster (Depart-ment of Haematology, NHS Blood and Transplant) for phlebotomy and sample collection/processing, Joana Batista (Department of Haematology, NHS Blood and Transplant) for assistance with patient visits to Cambridge and sample processing, Daniel Duarte (NIHR BioResource) for DNA sample preparation of ThromboGenomics samples, Lisa Skeates and Luis Campos (Hematopathology and Oncology Diagnostic Service [HODS], Cambridge University Hospitals NHS Trust) for preparation of bone marrow trephine specimens, Pedro Martin-Cabrera for providing control bone marrow trephine specimens (HODS, Cambridge University Hospitals NHS Trust), Daniel Cutler (University College London) for review of electron microscopy images, James Thaventhiran (University of Cambridge) for evaluation of CD4-lymphocyte data, and Paul Sims for checking the manuscript for accuracy of spelling, punctuation, and grammar.

M.C.S and J.H.C. are supported by MRC Clinical Research Training Fellowships (MR/R002363/1 and MR/P02002X/1). L.M. is supported by a PhD Studentship grant from the Rosetrees Trust. T.K.B was supported by NHS Blood and Transplant and the British Society for Haematology. D.S. was in part funded by an Isaac Newton Trust/Wellcome Trust Institutional Strategic Support Fund fellowship to M.F.. L.K. and A.S. acknowledge support by the Ministerium für Kultur und Wissenschaft des Landes Nordrhein-Westfalen, the Regierende Bürgermeister von Berlin - inkl. Wissenschaft und Forschung, and the Bundesministerium für Bildung und Forschung. L.B. was supported by a fellowship from Fondazione Umberto Veronesi. P.G. was supported by a Telethon Foundation Grant (GGP15063). K.F. is supported by the Research Council of the KULeuven (C14/19/096) and by unrestricted grants of Swedish Orphan Biovit-rum AB, Bayer and CSL Behring. M.F. is supported by the British Heart Foundation (FS/18/53/33863).

This work was performed using resources provided by the Cambridge Service for Data Driven Discovery (CSD3) operated by the University of Cambridge Research Computing Service, provided by Dell EMC and Intel using Tier-2 funding from the Engineering and Physical Sciences Research Council (EP/P020259/1), and DiRAC funding from the Science and Technology Facilities Council.

We thank NIHR BioResource volunteers for their participation, and gratefully acknowledge NIHR BioResource centers, NHS Trusts and staff for their contribution. We thank the National Institute for Health Research and NHS Blood and Transplant. The views expressed are those of the author(s) and not necessarily those of the NHS, the NIHR, or the Department of Health and Social Care in England.

## Authorship contributions

M.C.S., L.M., J.H.C., W.H.O., and J.A.G. wrote the manuscript. M.C.S., L.M., J.H.C., T.K.B., K.M., C.L-B., M-C.A., W.F.B., L.B., E.C., E.DC., K.G., A.G., P.G., D.H., M-F.H., A.M.K., R.K., S.LQ., T.L., E.B.L., A.D.M., S.M., D.N., S.P., S.J.P., J.P., G.M.P., M-C.P., M.S., H.S., S.S., O.S-S., R.C.T., J.K.M.W., B.Z., K.F., P.N., and R.F recruited participants into the study and collected phenotype data. M.C.S., L.M., J.H.C., T.K.B., L.K., F.S.B., S.F., R.D.S., R.M., H.M., R.R., J.C.S., W.N.E., and J.A.G. handled and processed samples and performed experiments. M.C.S., L.M., J.H.C., K.M., D.S., L.K., F.S.B., D.G., D.L., A.R-R., W.J.A., S.V.V.D., L.G., E.T., T.W.K., K.D., W.N.E., and J.A.G. analyzed data. E.T., A.D.W., A.S., K.D., W.N.E., M.F., W.H.O., R.F., and J.A.G. provided oversight of recruitment processes and analysis pipelines. W.H.O. and J.A.G. supervised the project.

A complete list of the members of the NIHR BioResource is provided in Supp. Table 4.

## References

1. Albers CA, Cvejic A, Favier R, et al. Exome sequencing identifies NBEAL2 as the causative gene for gray platelet syndrome. Nat Genet. 2011;43(8):735–737.

2. Gunay-Aygun M, Falik-Zaccai TC, Vilboux T, et al. NBEAL2 is mutated in gray platelet syndrome and is required for biogenesis of platelet alpha-granules. Nat Genet. 2011;43(8):732–734.

3. Kahr WH, Hinckley J, Li L, et al. Mutations in NBEAL2, encoding a BEACH protein, cause gray platelet syndrome. Nat Genet. 2011;43(8):738–740.

4. Lo RW, Li L, Leung R, Pluthero FG, Kahr WHA. NBEAL2 (Neurobeachin-Like 2) Is Required for Retention of Cargo Proteins by alpha-Granules During Their Production by Megakaryocytes. Arterioscler Thromb Vasc Biol. 2018;38(10): 2435–2447.

5. Mayer L, Jasztal M, Pardo M, et al. Nbeal2 interacts with Dock7, Sec16a, and Vac14. Blood. 2018;131(9):1000–1011.

6. Deppermann C, Cherpokova D, Nurden P, et al. Gray platelet syndrome and defective thrombo-inflammation in Nbeal2-deficient mice. J Clin Invest. 2013;123(8): 3331–42.

7. Guerrero JA, Bennett C, van der Weyden L, et al. Gray platelet syndrome: proinflammatory megakaryocytes and alpha-granule loss cause myelofibrosis and confer metastasis resistance in mice. Blood. 2014;124(24):3624–3635.

8. Kahr WH, Lo RW, Li L, et al. Abnormal megakaryocyte development and platelet function in Nbeal2-/-mice. Blood. 2013;122(19):3349–3358.

9. Claushuis TAM, de Stoppelaar SF, de Vos AF, et al. Nbeal2 Deficiency Increases Organ Damage but Does Not Affect Host Defense During Gram-Negative Pneumonia-Derived Sepsis. Arterioscler Thromb Vasc Biol. 2018;38(8):1772–84

10. Sowerby JM, Thomas DC, Clare S, et al. NBEAL2 is required for neutrophil and NK cell function and pathogen defense. J Clin Invest. 2017;127(9):3521–3526.

11. Drube S, Grimlowski R, Deppermann C, et al. The Neurobeachin-like 2 Protein Regulates Mast Cell Homeostasis. J Immunol. 2017;199(8):2948–2957.

12. Chedani H, Dupuy E, Masse JM, Cramer EM. Neutrophil secretory defect in the gray platelet syndrome: a new case. Platelets. 2006;17(1):14–19.

13. Drouin A, Favier R, Masse JM, et al. Newly recognized cellular abnormalities in the gray platelet syndrome. Blood. 2001;98(5):1382–1391.

14. Kahr WH, Dror Y. Gray platelet syndrome: macrothrombocytopenia with deficient alpha-granules. Blood. 2012;120(13):2543.

15. Rensing-Ehl A, Pannicke U, Zimmermann SY, et al. Gray platelet syndrome can mimic autoimmune lymphoproliferative syndrome. Blood. 2015;126(16): 1967–1969.

16. Steinberg-Shemer O, Tamary H. Gray platelet syndrome mimicking atypical autoimmune lymphoproliferative syndrome: the key is in the blood smear. Blood. 2018;131(24):2737.

17. Gunay-Aygun M, Zivony-Elboum Y, Gumruk F, et al. Gray platelet syndrome: natural history of a large patient cohort and locus assignment to chromosome 3p. Blood. 2010;116(23):4990–5001.

18. White JG, Brunning RD. Neutrophils in the gray platelet syndrome. Platelets. 2004;15(5):333–340.

19. Downes K, Megy K, Duarte D, et al. Diagnostic high-throughput sequencing of 2,396 patients with bleeding, thrombotic and platelet disorders. Blood. 2019;134(23):2082–91.

20. Simeoni I, Stephens JC, Hu F, et al. A high-throughput sequencing test for diagnosing inherited bleeding, thrombotic, and platelet disorders. Blood. 2016;127(23):2791–2803.

21. The NIHR BioResource, on behalf of the 100,000 Genomes Project. Whole-genome sequencing of rare disease patients in a national healthcare system. Biorxiv. 2019.

22. Richards S, Aziz N, Bale S, et al. Standards and guidelines for the interpretation of sequence variants: a joint consensus recommendation of the American College of Medical Genetics and Genomics and the Association for Molecular Pathology. Genet Med. 2015;17(5):405–424.

23. Westbury SK, Turro E, Greene D, et al. Human phenotype ontology annotation and cluster analysis to unravel genetic defects in 707 cases with unexplained bleeding and platelet disorders. Genome Med. 2015;7(1):36.

24. Arber DA, Orazi A, Hasserjian R, et al. The 2016 revision to the World Health Organization classification of myeloid neoplasms and acute leukemia. Blood. 2016;127(20):2391–2405.

25. Lentaigne C, Greene D, Sivapalaratnam S, et al. Germline mutations in the transcription factor IKZF5 cause thrombocytopenia. Blood. 2019;134(23):2070–2081.

26. Perez-Riverol Y, Csordas A, Bai J, et al. The PRIDE database and related tools and resources in 2019: improving support for quantification data. Nucleic Acids Res. 2019;47(D1):D442–D450.

27. Karczewski KJ, Francioli LC, Tiao G, et al. Variation across 141,456 human exomes and genomes reveals the spectrum of loss-of-function intolerance across human protein-coding genes. Biorxiv. 2019.

28. Ashkenazy H, Abadi S, Martz E, et al. ConSurf 2016: an improved methodology to estimate and visualize evolutionary conservation in macromolecules. Nucleic Acids Res. 2016;44(W1):W344–350.

29. Cullinane AR, Schaffer AA, Huizing M. The BEACH is hot: a LYST of emerging roles for BEACH-domain containing proteins in human disease. Traffic. 2013;14(7):749–766.

30. Jogl G, Shen Y, Gebauer D, et al. Crystal structure of the BEACH domain reveals an unusual fold and extensive association with a novel PH domain. EMBO J. 2002;21(18):4785–4795.

31. Nurden AT, Nurden P. The gray platelet syndrome: clinical spectrum of the disease. Blood Rev. 2007;21(1):21–36.

32. Pluthero FG, Di Paola J, Carcao MD, Kahr WHA. NBEAL2 mutations and bleeding in patients with gray platelet syndrome. Platelets. 2018;29(6):632–635.

33. Favier R, Roussel X, Audia S, et al. Correction of Severe Myelofibrosis, Impaired Platelet Functions and Abnormalities in a Patient with Gray Platelet Syndrome Successfully Treated by Stem Cell Transplantation. Platelets. 2019:1–5.

34. Di Angelantonio E, Thompson SG, Kaptoge S, et al. Efficiency and safety of varying the frequency of whole blood donation (INTERVAL): a randomised trial of 45 000 donors. Lancet. 2017;390(10110):2360–2371.

35. Takeuchi A, Saito T. CD4 CTL, a Cytotoxic Subset of CD4(+) T Cells, Their Differentiation and Function. Front Immunol. 2017;8:194.

36. Rorvig S, Ostergaard O, Heegaard NH, Borregaard N. Proteome profiling of human neutrophil granule subsets, secretory vesicles, and cell membrane: correlation with transcriptome profiling of neutrophil precursors. J Leukoc Biol. 2013;94(4):711–721.

37. Grassi L, Izuogu O, Jorge N, et al. Cell type specific novel lincRNAs and circRNAs in the BLUEPRINT haematopoietic transcriptomes atlas. Biorxiv. 2019.

38. Lonsdale J, Thomas J, Salvatore M, et al. The genotype-tissue expression (GTEx) project. Nat Genet. 2013;45(6):580–585.

39. GTEx Consortium. Genetic effects on gene expression across human tissues. Nature. 2017;550(7675):204–213.

40. Larocca LM, Heller PG, Podda G, et al. Megakaryocytic emperipolesis and platelet function abnormalities in five patients with gray platelet syndrome. Platelets. 2015;26(8):751–757.

41. Cunin P, Bouslama R, Machlus KR, et al. Megakaryocyte emperipolesis mediates membrane transfer from intracytoplasmic neutrophils to platelets. Elife. 2019;8:e44031

42. Lee DH, Chen Y, Arunoday B, et al. Integrative Genomic Analysis and Functional Studies Reveal GP5, GRN, MPO and MCAM as Causal Protein Biomarkers for Platelet Traits. Biorxiv. 2019.

43. Rip J, Van Der Ploeg EK, Hendriks RW, Corneth OBJ. The Role of Bruton’s Tyrosine Kinase in Immune Cell Signaling and Systemic Autoimmunity. Crit Rev Immunol. 2018;38(1):17–62.

44. Xia S, Liu X, Cao X, Xu S. T-cell expression of Bruton’s tyrosine kinase promotes autoreactive T-cell activation and exacerbates aplastic anemia. Cell Mol Immunol. 2019.

45. Bottega R, Pecci A, De Candia E, et al. Correlation between platelet phenotype and NBEAL2 genotype in patients with congenital thrombocytopenia and alpha-granule deficiency. Haematologica. 2013;98(6):868–874.

